# Shifting trends in clinical parameters among dengue-infected patients in Bangladesh during the 2024 outbreak: A study based on demographic characteristics and IgG status

**DOI:** 10.1101/2025.06.23.25330131

**Authors:** Imrul Hasan Tipo, Imteaj Uddin Chowdhury, Sharaf Wasima Rahman, Nadia Islam, Arrafy Rahman, Samiha Ferdous Nuha, Mohammad Rafiqul Islam, A.N.M. Shahriar Zawad, Md. Mahfuz Ahmed, Tahsin Tassawar Chy, Mohammad Seraj Uddin, Prasun Barua, Md. Golam Kabir, Mamunur Rashid Mahib, Nishat Sultana, Mohammad Razuanul Hoque

**Affiliations:** Department of Biochemistry and Biotechnology, University of Science and Technology Chittagong, Chattogram, Bangladesh; Department of Biochemistry and Molecular Biology, University of Chittagong, Chattogram, Bangladesh; Department of Life Sciences, Independent University, Dhaka, Bangladesh; Department of Genetic Engineering and Biotechnology, University of Chittagong, Chattogram, Bangladesh; Department of Microbiology, University of Chittagong, Chattogram, Bangladesh; Department of Biochemistry, Army Medical College, Chattogram, Bangladesh; Department of Biochemistry and Molecular Biology, Noakhali Science and Technology University, Noakhali, Bangladesh

**Keywords:** Dengue, Clinical parameters, Platelet, NLR, Hematocrit, ALT, AST, IgG, Primary and Secondary infection

## Abstract

Dengue fever, a common mosquito-borne viral illness in tropical and subtropical areas, is endemic in over one hundred countries. Recent dengue outbreaks in Bangladesh exhibited higher morbidity and a notable change in signs and symptoms, making the disease pattern more complicated. Yet the inadequate knowledge regarding the shifting of clinical parameters leads to inaccurate diagnoses and constrained treatment options. This study sought to assess the changing patterns of clinicopathological markers across various stages of the disease. Data were collected from 211 (137 dengue-positive and 74 dengue-negative) hospitalized patients in Chattogram, Bangladesh during the 2024 dengue outbreak. The dengue-positive patients were categorized into groups according to age, gender, and IgG status (primary or secondary infection). We evaluated the parameters throughout five stages of the disease utilizing Prism GraphPad-10. In our study, older (>40 years) patients exhibited lower platelet counts over an extended period. Besides, a decrease in hematocrit and hemoglobin levels, along with an increased ESR (Erythrocyte Sedimentation Rate), NLR (Neutrophil-Lymphocyte Ratio), ALT (Alanine Transaminase), and AST (Aspartate Transaminase) was noted among older (>50 years) patients. In males and females, differences in the changing patterns of platelet count, hematocrit, ALT, and AST were observed. Significant differences in many indices between IgG-negative and IgG-positive patients have been noted. Compared to IgG-negative patients, IgG-positive patients exhibited lower platelet counts and higher ESR, NLR, AST, and ALT. Interestingly, hematocrit changing patterns were similar in IgG-positive and IgG-negative patients, while the pattern for NLR was substantially different. Initial NLR was high (>4) in IgG-positive patients, reducing significantly (p***=0.0001) at the following stage and lowering to 2 until the last stage, while IgG-negative patients maintained NLR around three. This research will help to elucidate the disease patterns of current dengue infections by providing significant insights into the changing clinical parameters during the period of the disease.

**Author Summary:** The recent dengue outbreak in Bangladesh demonstrated high morbidity and irregular signs and symptoms. However, there is a lack of sufficient studies illustrating the alterations in clinical parameters associated with disease progression, which are necessary for a comprehensive understanding of the disease patterns. This study evaluates the changing patterns of clinical parameters throughout the disease course in 137 dengue patients in Bangladesh. We observed a greater severity of the disease among elderly patients and a variation in the changing pattern of several clinical parameters between males and females. Significant deviations of platelet count, neutrophil to lymphocyte ratio, and liver enzyme level in blood were noted among dengue infected patients. Most importantly, IgG positive patients had a different pattern of disease compared to IgG negative patients. Individuals with IgG positivity exhibited a low platelet count, an elevated neutrophil to lymphocyte ratio and liver enzyme levels, and significant fluctuations in these parameters throughout the disease course, in contrast to IgG negative patients. These findings will enhance the understanding of the infection pattern of current dengue outbreak and guide treatment strategies considering the clinical parameters.

## Introduction

Dengue fever is a viral disease caused by the four serotypes of dengue virus (DENV1-4), the most widespread arbovirus (arthropod-borne virus), transmitted by mosquitoes of the genus *Aedes*, particularly *Aedes aegypti* [1,2]. This virus began to spread around three centuries ago through zoonotic transmission [3], and its prevalence has increased substantially over the last 50 years [1]. in the first 20 years of this century, dengue cases have increased 10-fold, from 0.5 million to 5.2 million people [4]. Every year, around 50–100 million people are infected by dengue [5], and this includes 500,000 cases of life-threatening dengue hemorrhagic fever/dengue shock syndrome (DHF/DSS) [1]. The WHO has listed Dengue as one of the top ten global health threats in 2019 [4]

The general clinical characteristics of dengue fever include headache, fever with headache, retro-orbital pain, myalgia, arthralgia, intense general malaise, osteoarticular pain with or without rash, leukopenia, and some kind of bleeding [1,2]. As it progresses to DHF, severe symptoms may appear, such as vascular permeability (plasma leakage), thrombocytopenia, and hemorrhagic manifestations, which can lead to large hemorrhages in the gastrointestinal tract, resulting in DSS [1,2]. Despite being antigenically homogeneous, the immunogenicity of the four DENV serotypes differs highly [6], which increases susceptibility to the disease up to four different times[7]. Besides, concurrent infections by distinct serotypes cause severe complications, including DHF and DSS [8]. Currently, DENV infection is highly prevalent in Africa, the Eastern Mediterranean, the Americas, Southeast Asia, and the Western Pacific. The rise in cases in 2020 has made DENV a severe virus-caused disease after COVID-19, according to a recent WHO report, with the highest number of cases reported in countries like the Philippines, Vietnam, India, Colombia, and Brazil [9].

Bangladesh is also one of the vulnerable countries to DENV outbreaks due to its geographical position in the tropical and subtropical regions, like other Southeast Asian countries, which makes it an ideal habitat for DENV carrier Aedes mosquitoes[10]. Although dengue was first discovered in Bangladesh in 1964, the life-threatening DHF did not appear until 2000. In Bangladesh, the infection of dengue rose drastically in the last 5 years. The dengue outbreaks in 2019 surpassed all previous records with more than 0.1 million reported cases. The number remained significantly high in 2022 because of heavy rainfall, high humidity, and an extended rainy season [10]. The country has experienced the most catastrophic outbreak of dengue fever to date in 2023, affecting 0.32 million people with 1705 death cases followed by 0.1 million cases in 2024 [11]. During the outbreak, the infection rate was higher for males, while females had a higher mortality rate [12]. In addition, the morbidity and mortality rate greatly varied among different age groups, where older people accounted for a higher risk of infection and death [12].

Over the last decades, variations in the prevalence of different DENV serotypes have been observed, complicating the infection patterns and signs and symptoms that have changed significantly over the period [13]. Hence, there is a critical need to identify reliable pathological markers that can aid in the early diagnosis, accurate prognosis, and effective treatment of dengue cases, particularly in regions like Chattogram with a high disease burden [14].

In recent years, advances in medical research have highlighted the potential of various pathological markers as indicators of disease progression and severity in dengue. These markers encompass a range of hematological, biochemical, and immunological parameters that reflect the underlying pathophysiological processes associated with the infection. Parameters such as platelet count and platelet indices, hematocrit, liver function enzymes, and immune cell profiles have shown promise in providing valuable insights into the disease’s trajectory and clinical outcomes [15]. However, the applicability of these pathological markers in the context of Chattogram’s dengue landscape remains relatively unexplored. Factors such as regional variations in viral strains, host genetic factors, and healthcare infrastructure can significantly influence the diagnostic and prognostic utility of these markers.

The goal of this research is to bridge this knowledge gap by conducting a rigorous study to evaluate the clinical applicability of pathological markers in the diagnosis, prognosis, and treatment of dengue in Chattogram, Bangladesh. By systematically analyzing a diverse patient population, encompassing both mild and severe cases, this study seeks to identify key pathological markers that can aid healthcare practitioners in making uniform decisions regarding patient management. Ultimately, the insights garnered from this research endeavor may contribute significantly to the enhancement of dengue care strategies not only in Chattogram but also nationwide and inform broader efforts aimed at mitigating the impact of this infectious disease.

## Materials and methods

### Ethical consideration

This study was reviewed and approved by the ethical committee of the University of Chittagong (approval no: AERB-FBSCU-20240711-(1)). All the participants were informed of the study’s purpose, and formal consent was taken. Consent was taken from the guardian in the case of child patients. The confidentiality and anonymity of the patients were strictly maintained

### Study area and data collection

This cohort study was conducted on 211 hospitalized patients during the period from July 2024 to October 2024 at the Parkview Hospital, Chattogram, Bangladesh. A total of 137 patients having NS1 positive and/or IgM positive were used in this study, as well as 74 patients having similar symptoms, but both NS1 and IgM negative were used as a control group. NS1 and/or IgM positive hospitalized patients of all ages and genders were included in this study. Patients with malaria, blood disorders, urinary tract infections, and immune disorders and those who did not give consent for the study were excluded. Participants were divided into different groups based on age, gender, and presence of IgG. Data was analyzed for up to 5 stages of disease during hospitalization period. Hematological analysis, including platelet count, PDW (Platelet Distribution Width), MPV (Mean Platelet Volume), hemoglobin, hematocrit, leukocyte count, and liver function enzyme (ALT, AST) tests were performed in the diagnostic laboratory of the Parkview Hospital utilizing their standard protocols and instruments.

### Statistical analysis

The statistical analysis of data was conducted using the software - GraphPad Prism 10.3.1 (509) version. Initially, frequency analysis was conducted, followed by a t-test to check if a significant difference (p < 0.05) exists between the two groups. The Pearson correlation was carried out to investigate the correlation between groups.

## Results

### Demographic features of the study population

In this study, we have compared several clinical parameters at the initial stage of the disease and analyzed the changing patterns of them throughout the 5 stages of the disease among different groups of dengue-positive patients. A total of 137 people were used in the cohort of the disease group, and 74 non-dengue febrile illness patients having similar symptoms were used as a control group. Among 137 dengue-positive patients, 65% (89) had NS1 positive, 41.6% (57) had IgM and/or IgG positive, and 6.6% (9) had both NS1 and IgM/IgG positive. Among the disease population, 73 patients performed IgG tests where 33 (45%) had IgG positive and 40 (55%) had IgG negative results. In the disease group of 137 individuals, 51% (70) were males and 49% (67) were females and in the control group of 74 individuals, 49% (36) were males and 51% (38) were females. Based on age, seven groups were made from child to older, as shown in Fig 1.

**Fig 1.**
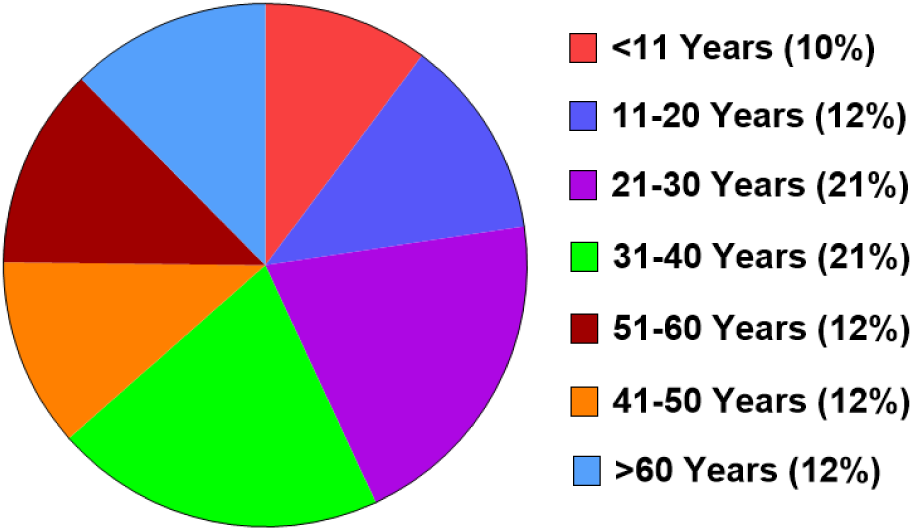
Distribution of dengue cases among different age groups (n=137)

### Comparison of Platelet count and platelet indices among different groups

#### Variation of Platelet count and platelet indices among different groups at the initial stage of disease

Total Platelet number greatly decreased across the groups of different ages compared to dengue negative patients (Fig 2A). Among them, children (<11 years) had a higher number of platelets than others, and adults (>41 years) had the least number of platelets. We also observed significant decrease (p<0.0001) of platelet count in both male and female patients compared to non-dengue individuals (Fig 2B). As IgG could be a good marker of secondary infection, we have compared the platelet number between IgG-positive and IgG-negative dengue patient groups as well. As predicted, IgG-positive individuals had a lower number of platelets than IgG-negative dengue patients, indicating more severe disease conditions during secondary infection (Fig 2C).

**Fig 2.**
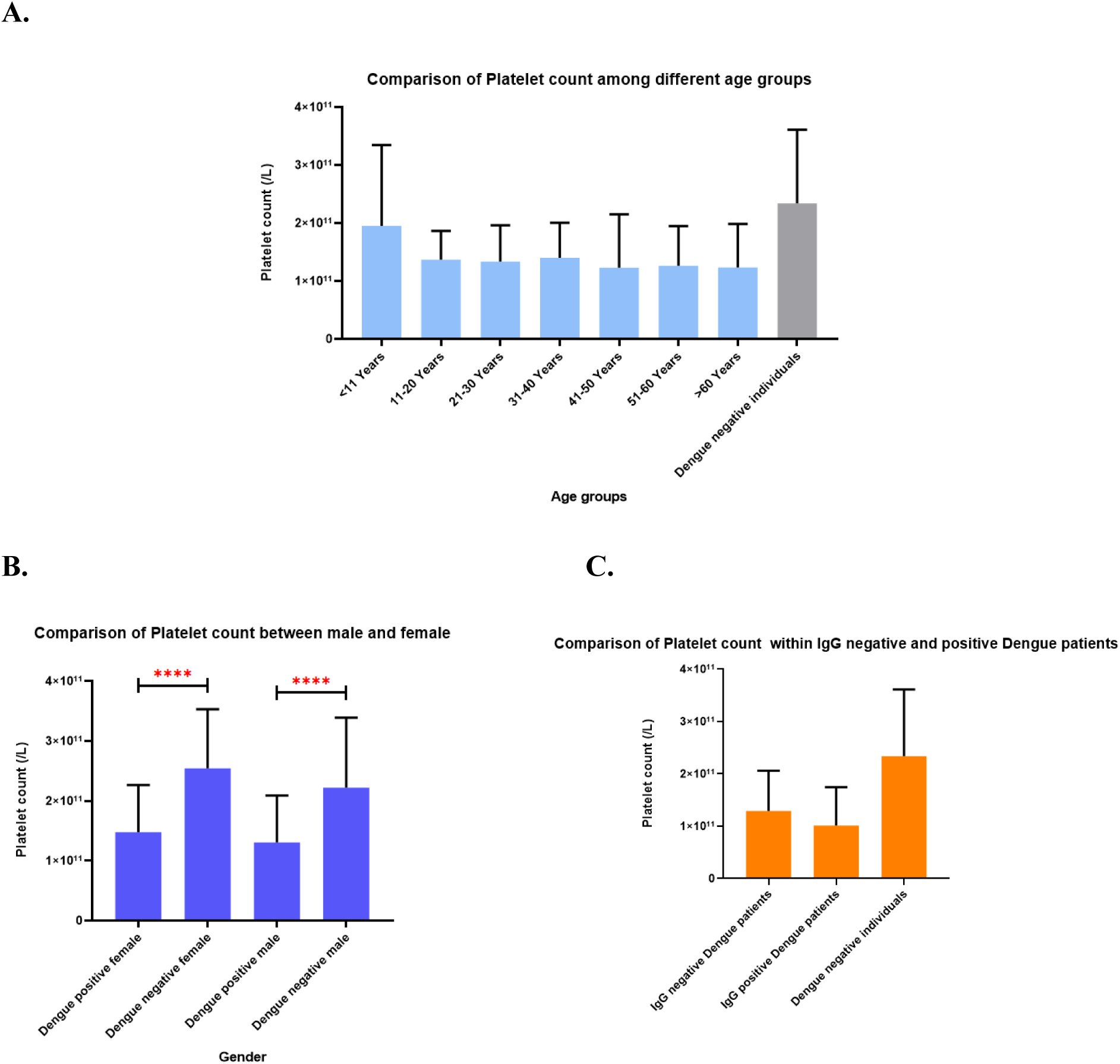
Comparison of platelet counts at the initial stage of disease among different (A) Age groups, (B) gender, and (C) IgG (+/-) groups.

Platelet indices, including platelet distribution width (PDW) and mean platelet volume (MPV), are also useful parameters during dengue infection [16]. PDW reflects the uniformity of platelet size, whereas MPV reflects the average size of platelets. We did not get significant correlations of PDW and MPV among different age groups, gender, and IgG (+/-) patients (S1 and S2 Fig).

#### Changing pattern of platelet numbers throughout the disease period

The platelet count of the dengue patients was evaluated at 5 different stages of disease. In most of the cases, platelet numbers decreased till stage 3 or 4, indicating the severe phase of illness, and then increased at stage 5, indicating recovery. Interestingly, we got a significant decline of platelet numbers between stage-1 to stage-2 in age group-1(p***=0.0001), group-2(p**=0.0027), group-4(p***=0.0003), and group-7(p**=0.0025) (Fig 3A). However, age groups -3,5 and 6 did not show a significant correlation among different disease stages.

**Fig 3.**
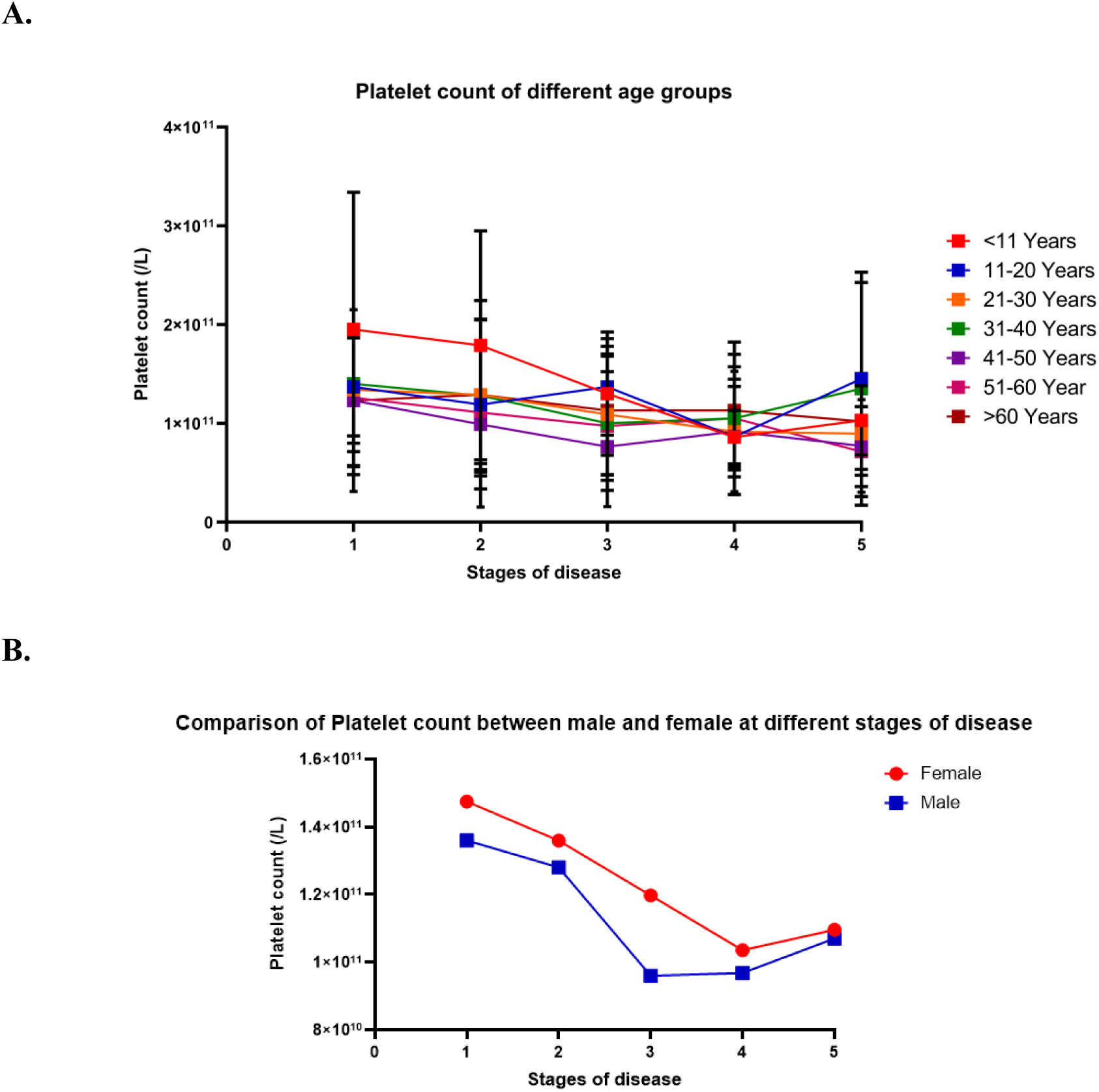
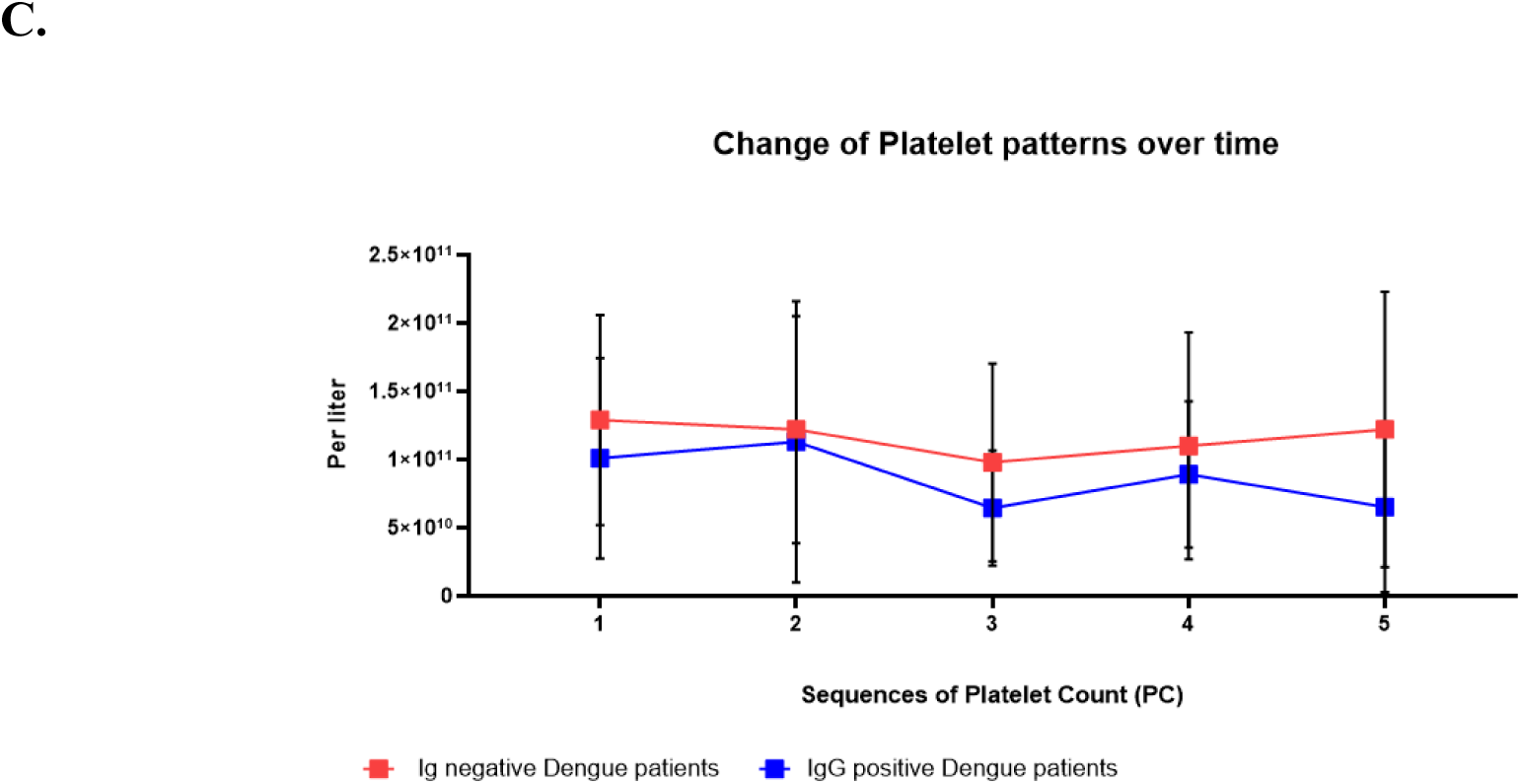
Patterns of platelet count changed with progression of disease among different (A) Age groups, (B) Gender groups, (C) IgG (+/-) groups.

The platelet count curve declined from stage-1 to stage-4 in females and stage-1 to stage-3 in males, and both recovered at stage 5 (Fig 3B). Again, we got a highly significant correlation of decreasing the numbers in stage-1 and stage-2 of both females (p<0.0001) and males (p<0.0001). But, in the t-test, no significant correlation between males and females was observed.

In IgG-negative dengue patients, the platelet count decreased from stage 1 to stage 2 (p****<0.0001) and stage-2 to Stage 3 (p**=0.016) significantly. Surprisingly, the platelet count graph was very unusual among IgG positive patients. The number increased from stage-1 to stage-2 (p****<0.0001) and decreased (p*=0.0286) from stage-2 to stage-3, followed by further rising (p****=0.0001) during stage-3 to stage-4 among dengue IgG positive individuals. We observed significantly lower platelets among IgG-positive patients at the middle (p=0.0218) and the last (p=0.059) stage of the disease compared to IgG-negative patients (Fig 3C).

### Evaluation of hematocrit (Ht), hemoglobin, and ESR level

#### Ht level of patients across different stages of the disease

Hematocrit level represents the proportion of RBCs in blood. A low Ht level indicates anemia, where the level increases in polycythemia [17]. During the initial days (Stage1 to stage-2) of infection, the Ht level declined among children (p**=0.0072) and adolescents (p***=0.0004), whereas the level rose significantly among older groups - group-5 (p*=0.0353) and group-6 (p****<0.0001). The level remained static among younger (21-30 years) throughout the disease periods (Fig 4A). In addition, males had a lower Ht level than females (Fig 4B), and both IgG+ and IgG-patients had similar patterns of Ht levels during infection periods (Fig 4C).

**Fig 4.**
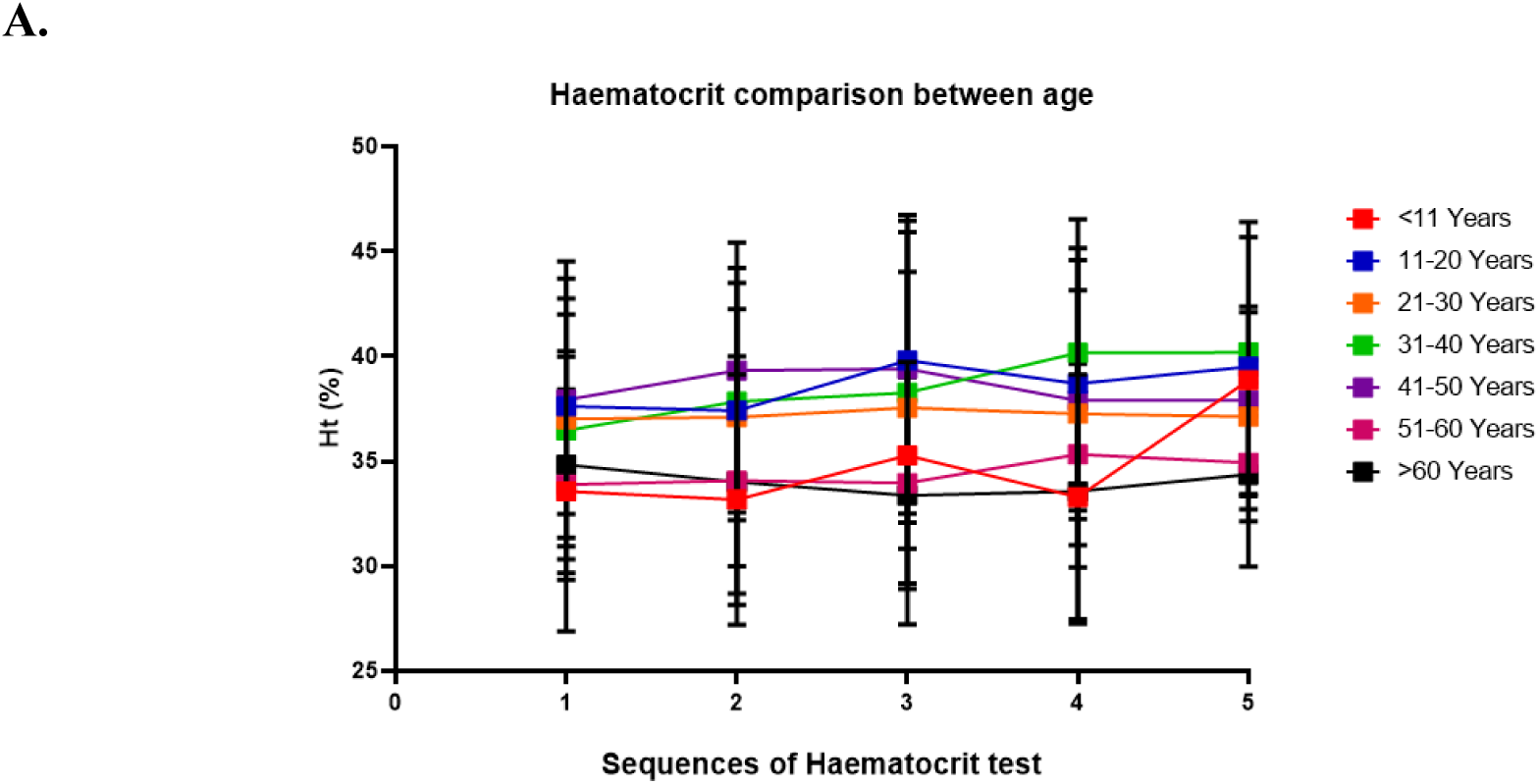
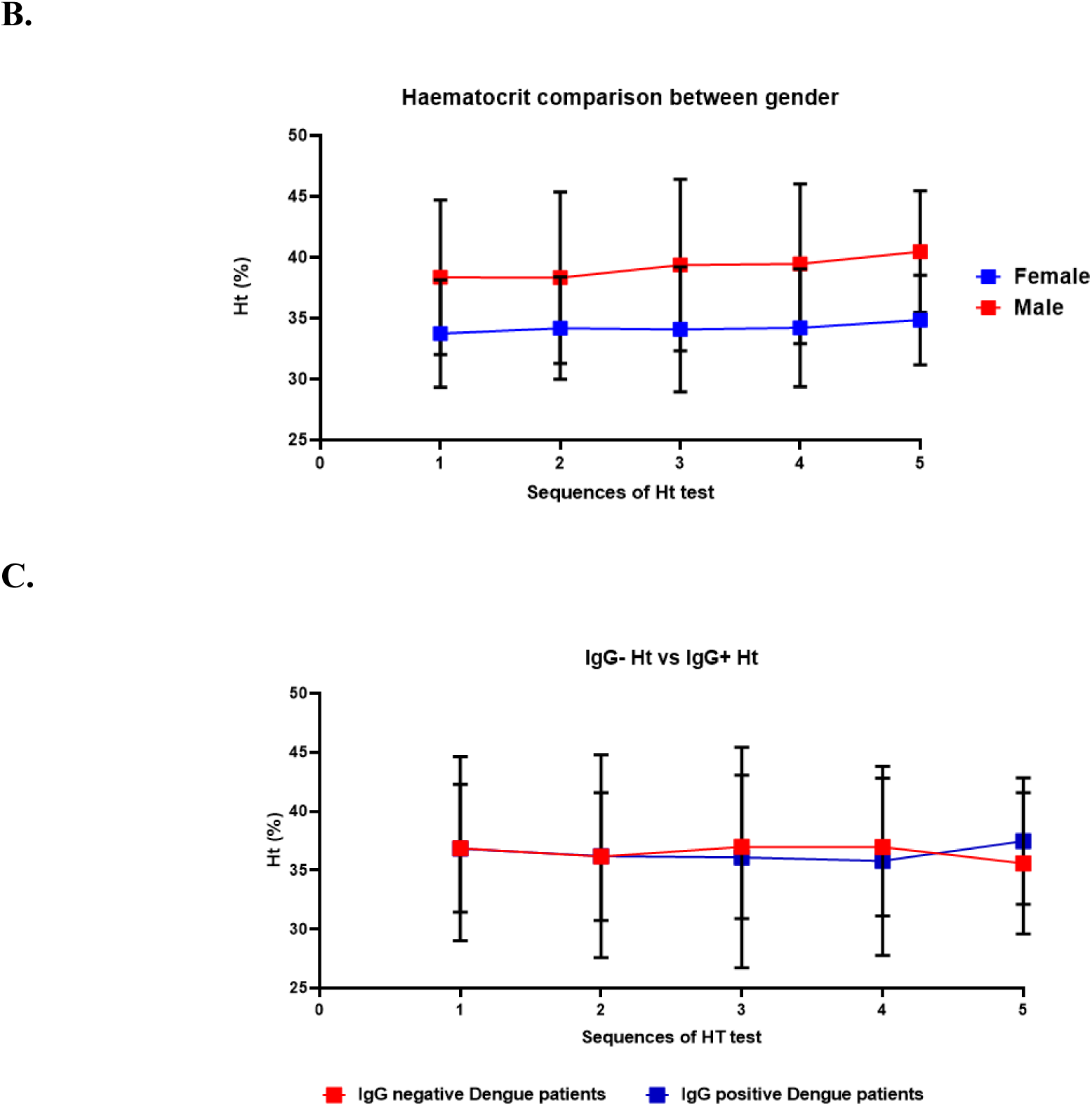
Comparison of Ht changing patterns during different disease stages among distinct groups of (A) Age, (B) Gender, and (C) IgG.

#### Comparison of hemoglobin level among different groups

The hemoglobin concentration (gm/dl) was compared among different age groups. This comparison has also been carried out between genders as well as IgG-positive and IgG-negative dengue patients. The hemoglobin concentration has ranged between 10 mg/dl to 15 mg/dl among different age groups with no significant difference (Fig 5A). Although hemoglobin concentrations are visually the same in the case of dengue-positive and dengue-negative females, a significant difference (p = 0.023) has been found between dengue-positive and dengue-negative males (Fig 5B). No significant difference has been observed between the hemoglobin concentrations of IgG-positive and IgG-negative patients (Fig 5C).

**Fig 5.**
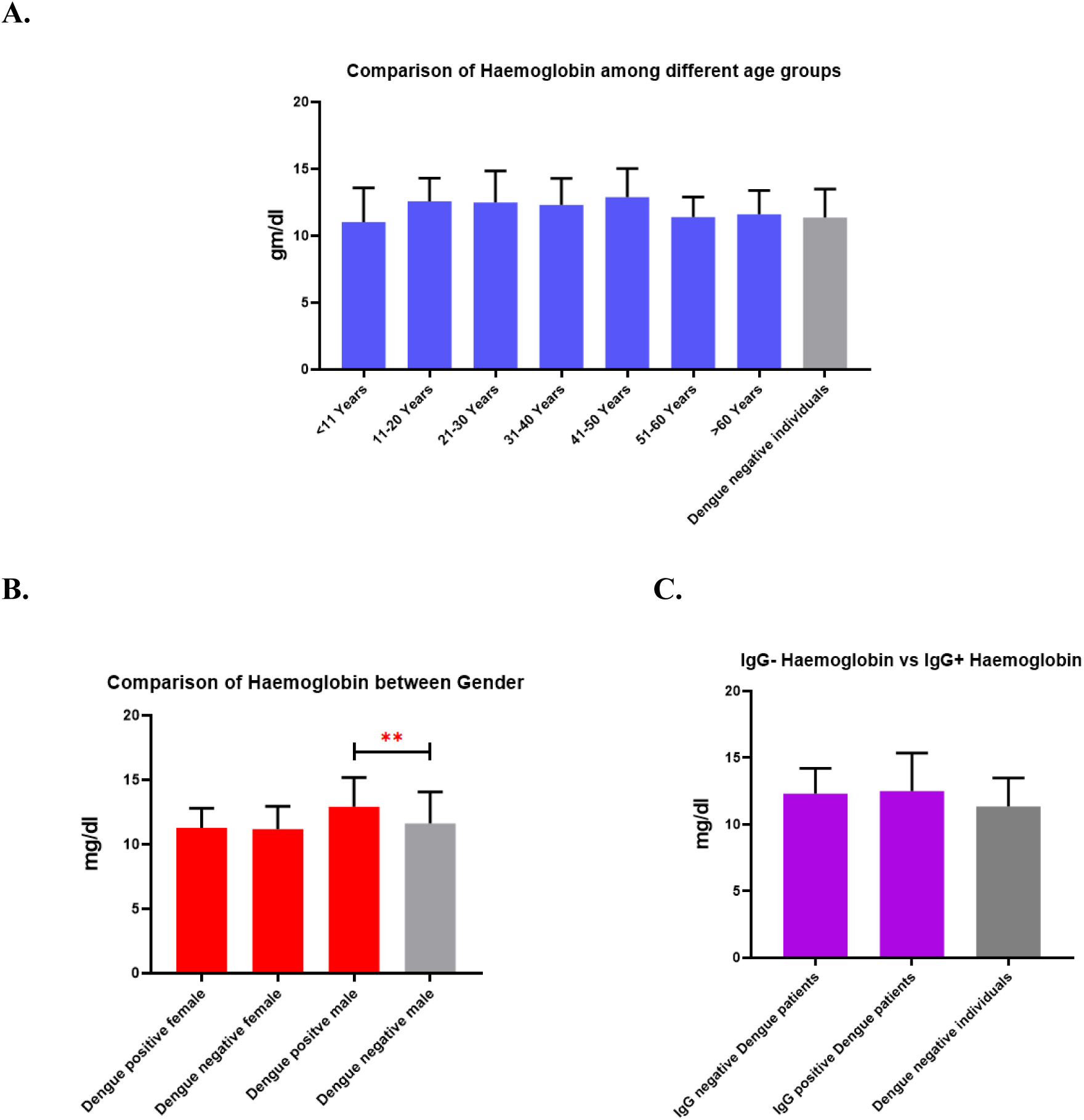
Hemoglobin levels in different (A) Age, (B) Gender, and (C) IgG groups at the primary stage of disease.

#### Comparison of ESR level

The Erythrocyte Sedimentation Rate (ESR) varied throughout the age groups, with the lowest ESR (∼12 mm/hour) found in patients between 11 and 20 years and patients aged 51-60 years accounted for the highest ESR, which is >40 mm/hour (Fig 6A). The ESR of dengue-positive females (∼30 mm/hour) significantly lowered (P<0.001) from that of Dengue-negative females (∼55 mm/hour) (Fig 6B). The ESR of IgG-positive patients was higher than that of IgG-negative patients, but no significant difference was observed (Fig 6C).

**Fig 6.**
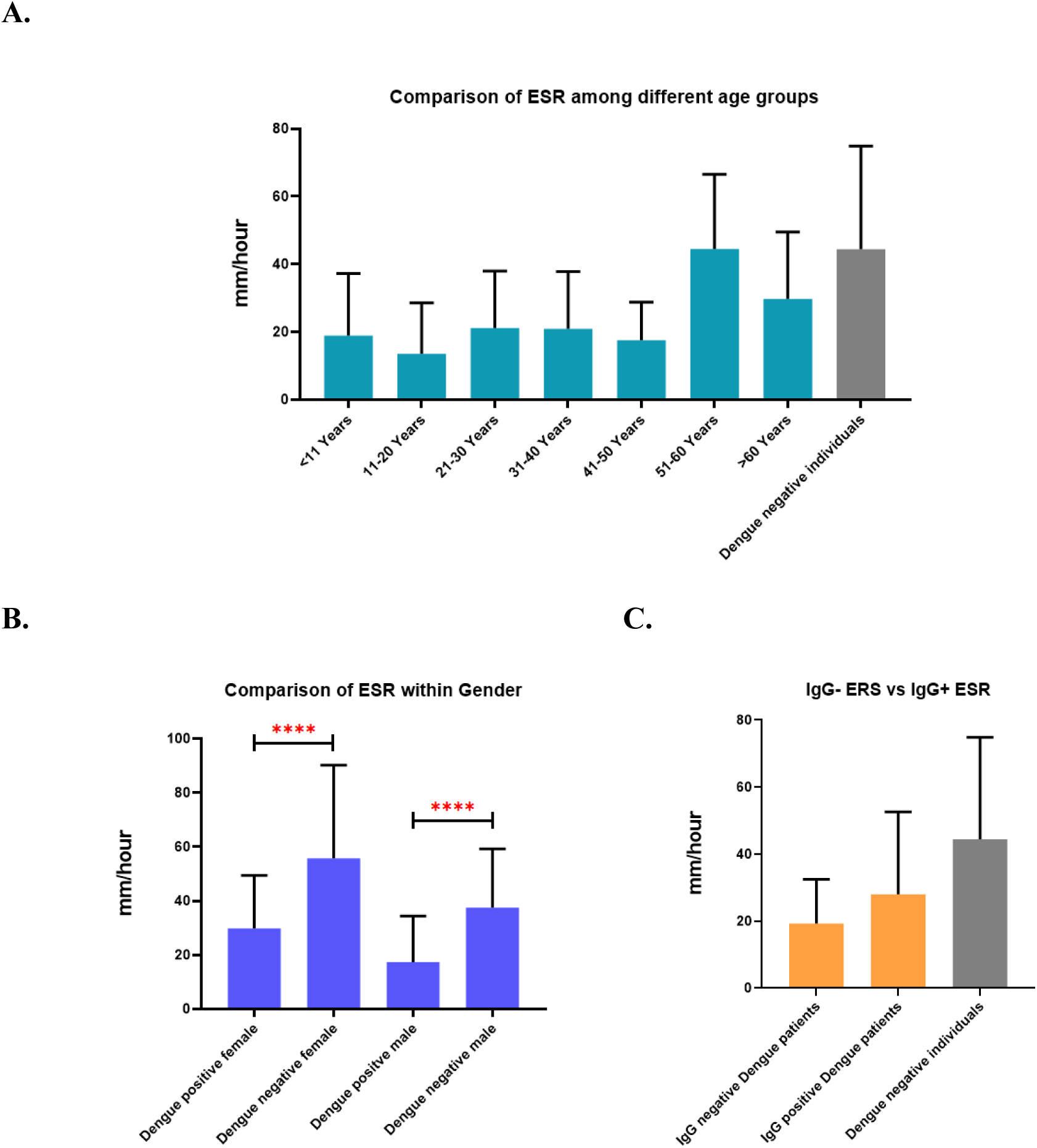
The level of ESR among the patients of different (A) age, (B) Gender, and (C) IgG groups.

### Analysis of neutrophil to lymphocyte ratio (NLR)

#### NLR value at the primary stage of disease among different groups

As the neutrophil-to-lymphocyte ratio (NLR) is positively correlated to inflammatory responses [18], we analyzed the ratio among different groups of patients. Initially, the NLR was around 5 in most of the age groups, and the lowest NLR was exhibited in the age group of 11-20 years (Fig 7A). The NLRs of male and female patients were very close to each other, whereas a slight difference has been observed in the NLRs of IgG-positive and IgG-negative patients (Fig 7B and 7C). Interestingly, the NLR ratio was higher among control group patients with febrile fever (Fig. 7).

**Fig 7.**
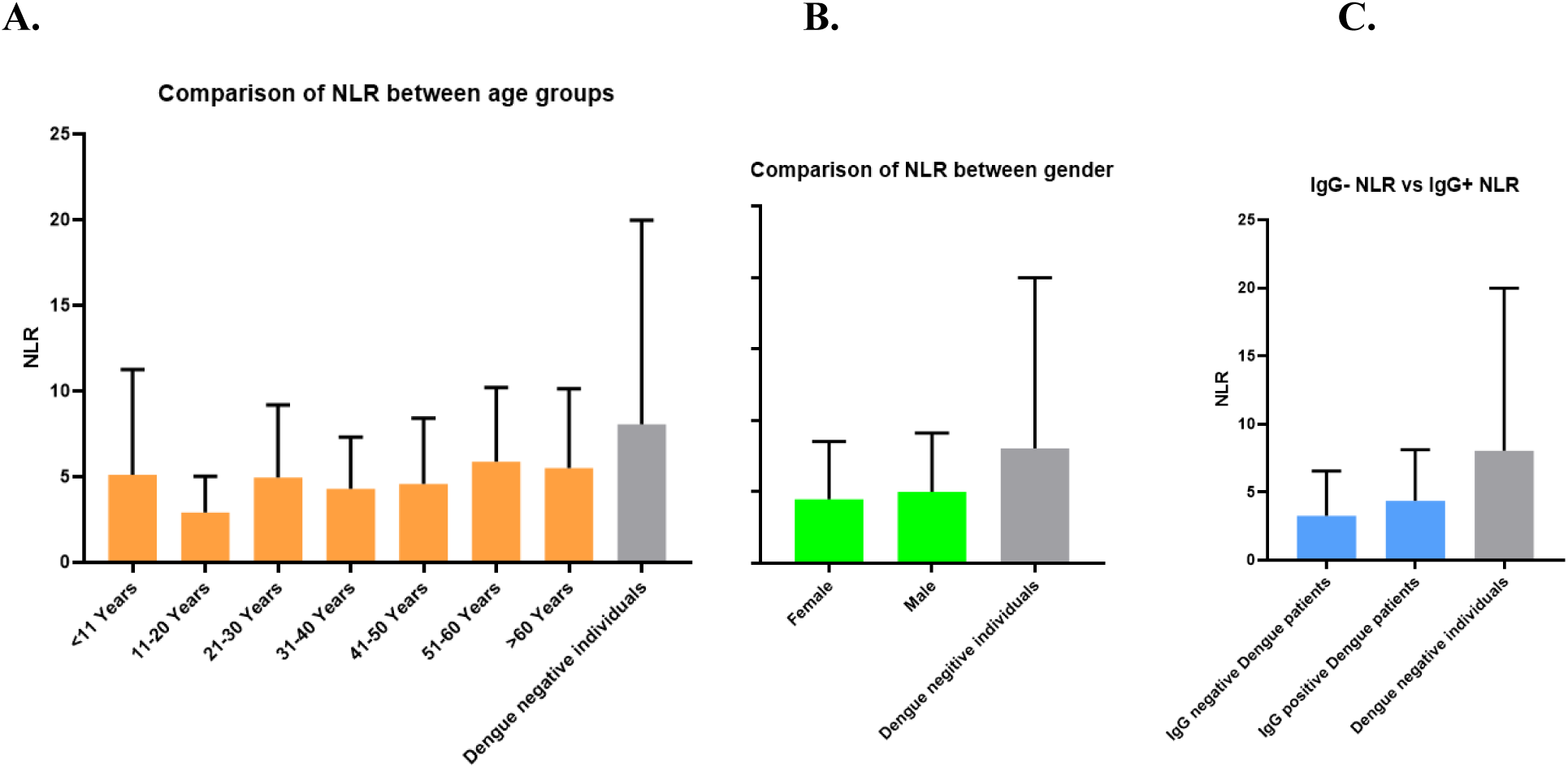
Comparison of NLR among different (A) Age, (B) Gender, and (C) IgG groups.

#### Changing patterns of NLR during disease progression

This study also evaluated the NLR level throughout the disease period. The NLR has decreased sharply from the initial stage to stage-3 in all age groups except the group of 21-30-year-old patients. A significant NLR fall was observed in the initial days (stage-1 to stage-2) among the patients aged 31-40 years (p***=0.0004) and the patients aged over 60 years (p**=0.0095). Among children (<11), NLRs were also significantly decreased (p**=0.0027) from stage 2 and stage 3 of the disease (Fig 8A). In both males and females, there was a sharp decrease (p = 0.0495 and 0.0123, respectively) in NLR from stage 1 to stage 2, followed by stage 3. After stage 3, the NLR value remained relatively constant (Fig 8B).

**Fig 8.**
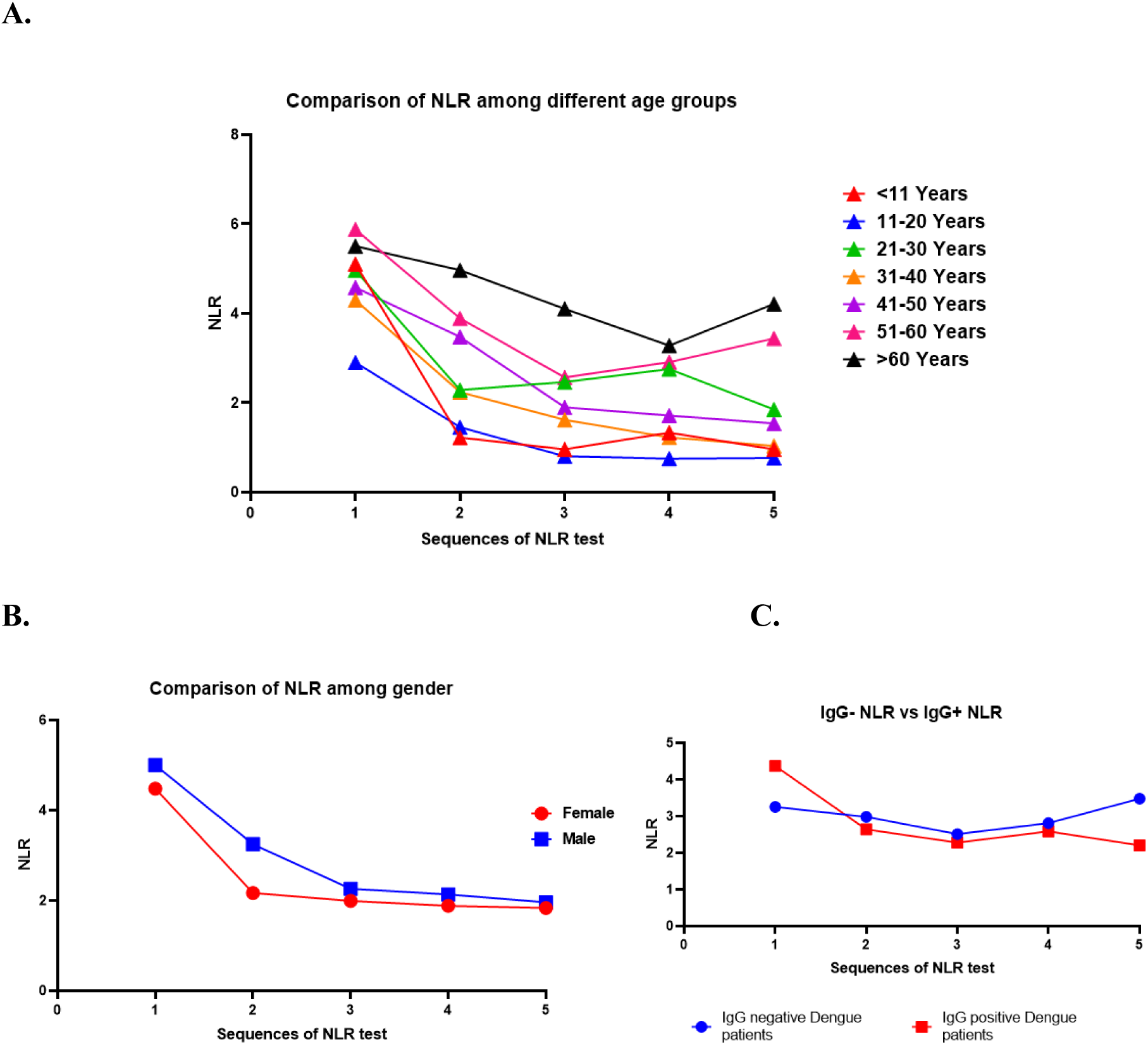
Fluctuation of NLR among different (A) Age, (B) Gender, and (C) IgG groups throughout the disease period.

A high difference in NLR changing patterns between IgG-positive and IgG-negative patients was observed. Initially, the NLR was higher among IgG-positive patients, indicating severe inflammation compared to IgG-negative individuals. Although the value decreased (p***=0.0001) sharply from 4.5 to 3 in the next step among IgG-positive patients, no significant change was observed among IgG-negative patients. Throughout the disease period, there was very little fluctuation in NLR value among IgG-negative patients. On the other hand, the value deviated significantly (particularly at the initial and last stage) among IgG-positive individuals (Fig 8C).

### Comparison of ALT and AST levels among different groups of patients

#### ALT level among different groups of patients

The ALT level rose greatly in all age groups of dengue-positive patients compared to the control group (Fig 9A). The concentration of ALT in male patients (∼650 u/l) was significantly higher (p= 0.036) than that of female patients (∼180 u/l) (Fig 9B). The ALT concentration of IgG-positive patients was higher than the ALT concentration of IgG-negative patients, but this difference was statistically non-significant (Fig 9C).

**Fig 9.**
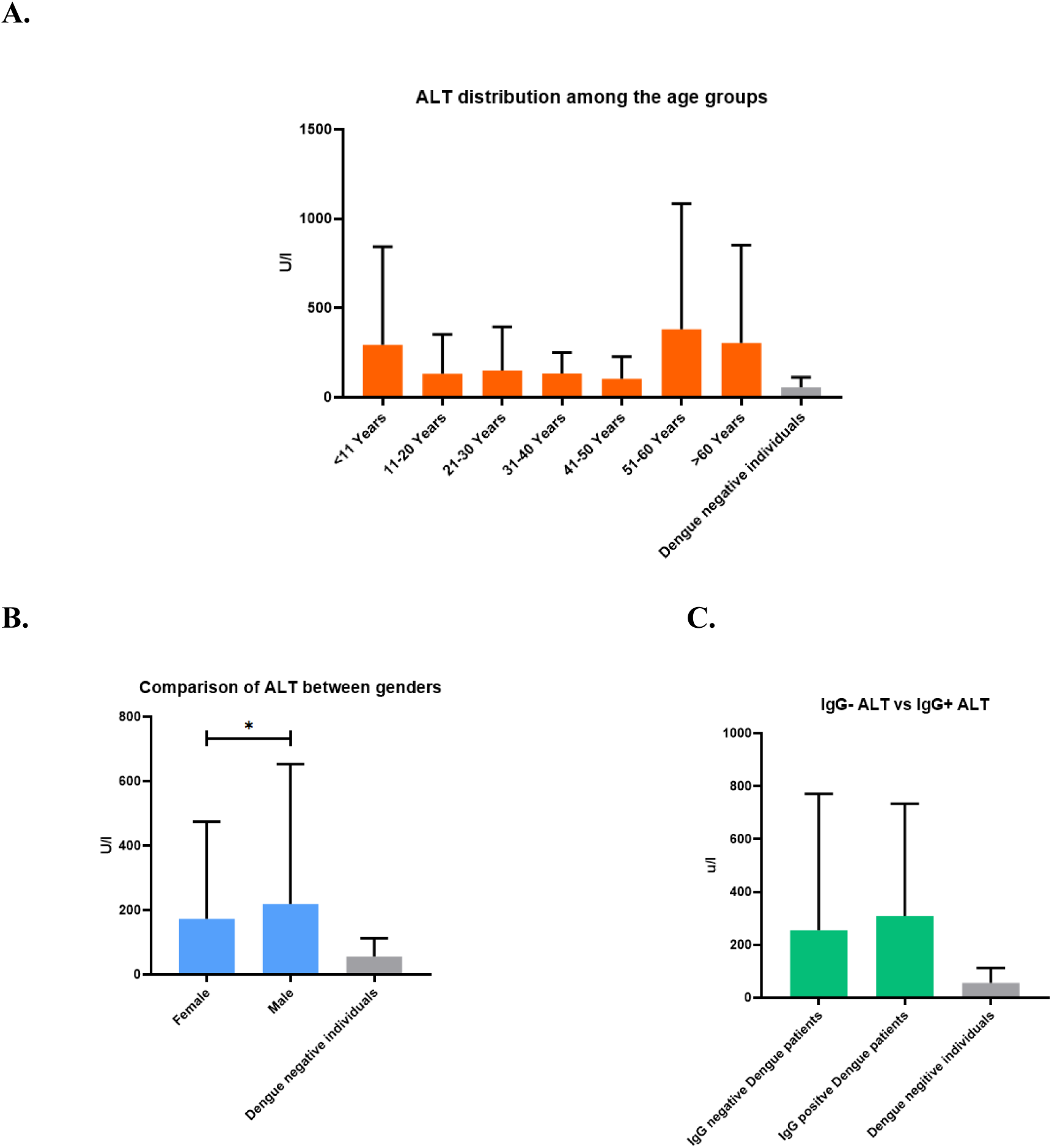
The ALT level among different (A) Age, (B) Gender, and (C) IgG groups.

#### AST level among different groups of patients

The children (<11 years) and olds (>60 years) displayed high AST concentration compared to others. Patients aged from 11 to 30 years, as well as 41 to 50 years, had the lowest concentrations of AST (Fig 10A). Male dengue patients had an AST concentration of ∼500 u/l, and female Dengue patients had over 500 u/l (Fig 10B). Surprisingly, the AST concentration of IgG-positive patients was 3 times more than that of IgG-negative patients (Fig 10C).

**Fig 10.**
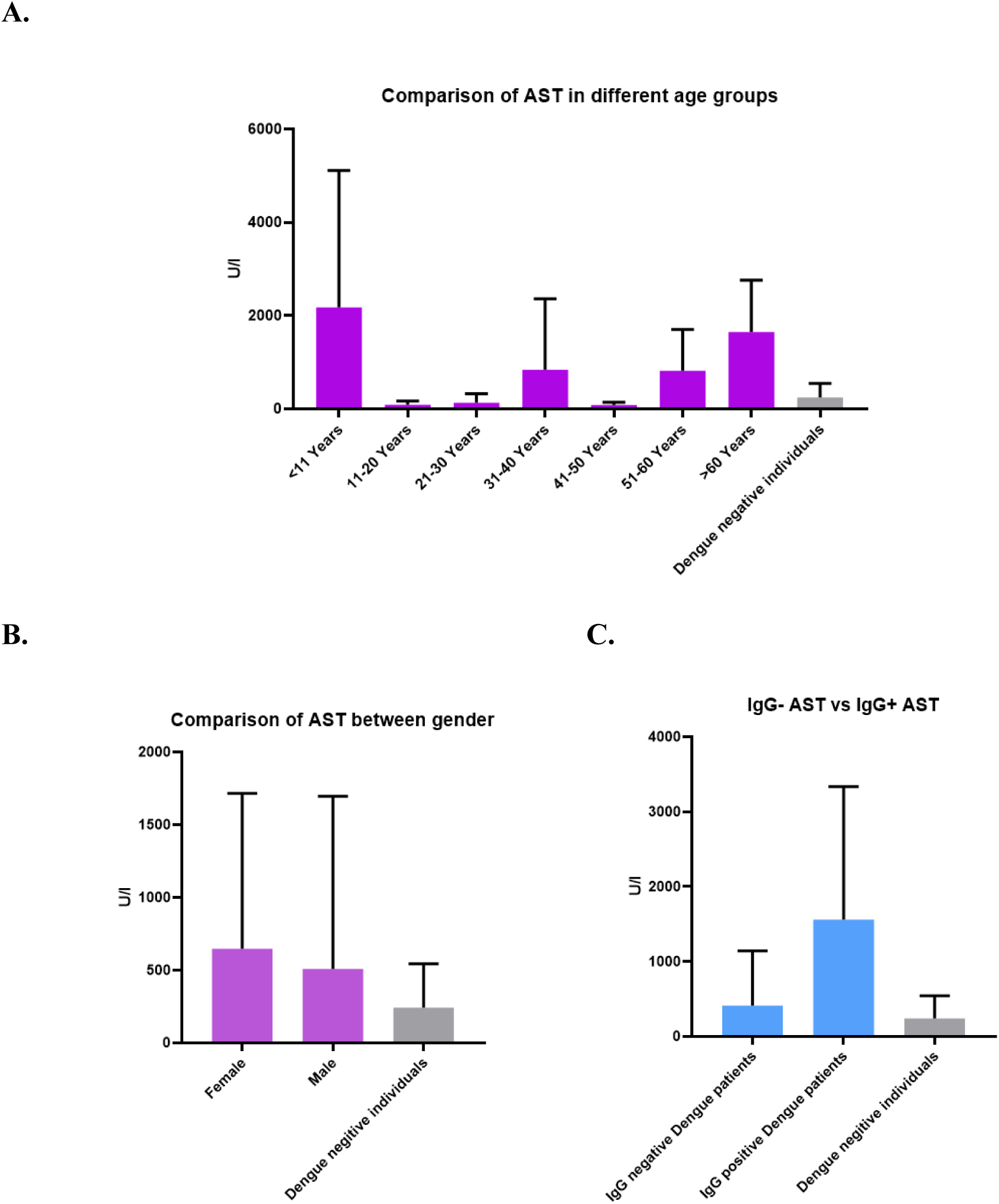
The AST level among different (A) Age, (B) Gender, and (C) IgG groups.

## Discussion

Dengue fever is a significant public health concern in many tropical and subtropical regions, including Bangladesh. Chattogram, a major port city in Bangladesh, has experienced recurrent outbreaks of dengue fever, leading to substantial morbidity and mortality. The lack of specific symptoms in the early stages of the disease often results in delayed diagnosis and initiation of treatment, which can exacerbate disease severity and complications. Accurate and timely diagnosis, prognosis, and treatment are critical for managing dengue cases effectively [19]

It is difficult to differentiate between Dengue fever and other mosquito-borne diseases as they share a wide range of similar clinical characteristics, particularly after illness onset. However, a number of markers are considered a potential indicator of severe dengue fever indicators of severe dengue fever, which may help in early diagnosis and monitoring disease progression [20]. This research aims to compare different hematological parameters related to clinical outcomes among different groups of dengue-positive patients at various stages of the disease in Chattogram, Bangladesh, in 2023. Platelet counts, Platelet indices (PDW & MPV), Hematocrit (HCT), Hemoglobin (Hb), Erythrocyte Sedimentation Rate (ESR), Neutrophil to Lymphocyte ratio (NLR) and Liver enzymes (ALT & AST) are used here as pathological markers to evaluate the immune and physiological changes in dengue infection, providing insights into disease severity and potential markers for monitoring patient recovery.

A total of 211 patients were included in the study, with 137 (70 Male & 67 Female) patients being dengue positive and 74 (36 Male & 38 Female) patients having non-dengue febrile illness showing similar symptoms were used as controls. Among the 137 dengue-positive patients, 65% (89) were NS1 positive, indicating early-stage infection, while 33 patients were IgG positive, demonstrating a secondary infection. During the first 3 days of fever onset, NS1 antigen is usually found to be relatively high [21], and NS1 positivity after 5 days of the fever indicates a high susceptibility to severe dengue [22]. NS1 is a highly precise viral marker for both primary and secondary infection, which makes it an extremely reliable parameter for the diagnosis of dengue [23]. The IgG is usually not detected in the early days of dengue primary infection. Instead, IgG can be found positive by the fourteenth day. On the other hand, during the early days of secondary infection, IgG levels rise significantly, surpassing IgM levels [24]. As a result, positive IgG results during the early stages of the disease could be a good marker for identifying secondary infection. Interestingly, primary infection with one serotype can lead to a secondary infection with a different serotype, resulting in severe dengue [25]. Thus, IgG as a marker can denote the severity of dengue infection.

In the study, platelet counts were found to be significantly reduced across all age groups in dengue-positive patients compared to controls, which aligns with the condition of thrombocytopenia (abnormally low level of platelets). According to [23], dengue-positive patients are more susceptible to thrombocytopenia than dengue-negative patients. In dengue patients, platelet counts can fall rapidly, and platelets less than 100,000/L increase the severity of the disease [26]. In our study, platelet levels sharply decreased from disease onset (stage 1) to severe illness (stage 3-4). However, patients showed recovery in stage 5, suggesting that platelets can act as an indicator for both disease severity and recovery. Among different age groups, adults (41+ years) exhibited the lowest platelet counts, compared to the children (0-11 years). This finding is consistent with the study by [20], which suggests that a low platelet count is an indicator for DHF in adult patients but not in pediatric patients. It can be due to age-related variance in immune response, as younger individuals usually have stronger innate immune responses.

Moreover, IgG-positive patients had lower platelet counts compared to the patients who were IgG-negative, suggesting that secondary infections might cause more severe immune responses, which can lead to more significant platelet destruction or consumption, possibly because of antibody-dependent enhancement (ADE) of dengue infection. A study conducted by [26] demonstrated that low platelet counts caused immunological responses like bone marrow suppression as well as liver and spleen-induced platelet destruction. Such findings emphasize the importance of monitoring platelet levels and IgG positivity, particularly in individuals who are at risk of secondary infection. However, thrombocytopenia can also be caused by some non-dengue febrile diseases, which indicates the necessity of using other dengue markers such as NS1, HCT, and NLR along with platelet counts.

Furthermore, the platelet indices, PDW (Platelet Distribution Width), and MPV (Mean Platelet Value) can act as indicators to evaluate the prognosis and understand platelet activation as well as turnover in dengue infection [16]. PDW measures platelet size variability, while MPV indicates the average platelet size. In our study, the values of MPV and PDW were slightly variable among dengue-positive patients and dengue-negative patients. The value either slightly elevated or declined in dengue-positive individuals. High MPV can cause peripheral destruction with ongoing thrombocytopenia, while Low MPV indicates bone marrow suppression. Moreover, PDW is associated with the value of MPV, that is, a high PDW value indicates a high MPV. PDW is increased in the presence of platelet anisocytosis (non-uniform platelet size), which can be caused by bone marrow disorder, recovery from thrombocytopenia, etc. [16]. While not all findings were statistically detailed, these indices contribute additional layers of insights into dengue diagnosis with hematological impact, especially when used alongside platelet counts.

Hematocrit (HCT) levels, which indicate the proportion of blood volume occupied by RBC, were found notably higher in our study in severe stages of dengue infection in specific age groups, pointing to hemoconcentration (reduced plasma volume with increased concentration of blood components), likely due to plasma leakage, that can lead to DHF or DSS if not diagnosed appropriately [16,27]. A decrease in HCT may indicate a recovery phase of the disease [16]. Increased HCT levels support clinical observations of fluid imbalance and dehydration in severe cases, making HCT a valuable and reliable parameter in evaluating dengue severity. Interestingly, while hemoglobin (Hb) levels did not show statistically significant changes across different stages or groups in our study, ESR values were generally elevated among dengue-positive patients. In severe cases, dengue can cause increased Hb levels temporarily due to hemoconcentration, which can also elevate HCT levels. However, the absence of significant findings for Hb in our study could indicate that most patients did not experience extreme plasma leakage or hemorrhage, suggesting hemoglobin is a less sensitive marker to demonstrate the severity of the disease. ESR measures inflammation in the body, and a faster rate indicates inflammation, infection, or immune response. In spite of varying statistical significance, ESR levels were generally high in dengue-positive patients in our study.

Increased ESR in dengue patients indicates an inflammatory response to viral infection. However, the lack of significant correlations across stages suggests that ESR might be a non-specific marker, meaning that while it reflects inflammation, it does not necessarily correlate with the clinical observations in dengue. While both Hb and ESR have roles in dengue infection, they may offer limited clinical reliability as a marker for the disease severity. These parameters should be interpreted alongside a broader hematological profile, including platelet counts, HCT, NLR, and liver enzyme levels, for the accurate diagnosis of the disease.

Neutrophil-to-Lymphocyte Ratio (NLR) is a marker of systemic inflammation, and when elevated, it may indicate a severe immune response [27]. It is calculated based on the ratio of neutrophil percentage (generally increased in response to infection and inflammation) to lymphocyte percentage (often depleted in viral infections like dengue) [28]. Increased NLR may indicate an intense inflammatory response, often seen in severe dengue, as the infection leads to an elevated neutrophil count while suppressing lymphocyte levels. Recently elevated neutrophil level with a high NLR ratio was found in the initial stage of the infection, and as the disease progressed from acute to severe phase, the ratio decreased due to declined neutrophil levels and increased lymphocyte percentage [28]. This finding demonstrated the importance of NLR as a marker to identify the disease in the initial stage. Our study found significant stage-wise changes in NLR, especially between the early stages (stages 1-2) of the disease in several groups. Stages 1 to 2 showed significant correlations in specific age groups, particularly groups 4 and 7, which indicated age-related immune responses in dengue, with certain age groups showing stronger or prolonged inflammatory responses compared to others. Our study did not show significant gender-based differences in NLR levels as the disease progressed, indicating that NLR reflects early inflammatory response in both genders. IgG-positive patients had elevated NLR, suggesting that those with secondary infections may experience heightened inflammatory responses, which can lead to severe clinical complications. Interestingly, throughout the disease stages of primary infection, the NLR value remains in an almost steady state, whereas in secondary infection, the ratio deviated significantly, indicating a remarkable change in neutrophil and lymphocyte numbers during secondary infection. Thus, in addition to indicating immune activation and inflammation, the differences in NLR trends between primary and secondary infections highlight the potential of NLR as an important marker for identifying primary and secondary dengue infection.

ALT (Alanine Aminotransferase) and AST (Aspartate Aminotransferase) enzymes are markers of liver function. Patients with DF and DHF have significantly higher ALT and AST levels compared to other patients with febrile illness [29], which indicates liver stress or injury during dengue. When liver cells are damaged or stressed, as can occur in viral infections like dengue, these enzymes are released into the blood circulation, leading to increased ALT and AST levels [30]. In our study, both ALT and AST were found elevated in dengue-positive patients in both genders than dengue-negative individuals, suggesting hepatic involvement. The increasing value varied in different age groups. Furthermore, increased levels of ALT and AST were also found in IgG-positive patients compared to IgG-negative patients, indicating a correlation between secondary infection and liver inflammation. In dengue, liver enzyme elevation can be mild to moderate in non-severe dengue but can reach significant levels in severe dengue patients [30]. In our study, patients were infected with non-severe dengue as the ALT and AST levels were not substantially higher. However, AST level was found to be higher than ALT, which can be due to AST’s presence not only in the liver but also in other organs, heart, muscles, and kidneys [30]. These elevated liver enzymes may serve as an early warning sign for severe dengue. Monitoring ALT and AST levels along with other markers such as platelet count and hematocrit, could help in the diagnosis of the disease to severe dengue. For instance, the combination of a high AST to ALT ratio with thrombocytopenia, high HCT, and elevated NLR could strengthen predictions of severe dengue, allowing for timely diagnosis and prognosis of the disease.

However, our study has some limitations. With a sample of 137 dengue-positive patients, the study may not evaluate the full variability in clinical presentations and a large sample size could improve statistical power. The findings may have been influenced by local factors, such as genetic variability, prevalent dengue serotype, and healthcare practices if the study was conducted within a specific region. Individual tracking of the patients over time might have allowed for a full exploration of how parameters change throughout the course of the illness. Factors like co-existing medical conditions and certain medications may have influenced some results of the non-specific markers like AST, ALT, and ESR since these markers can be elevated in various conditions beyond dengue.

The limitations do not outweigh the pros of this study for future implications in dengue-based research. Future studies should consider multicenter designs across various geographic regions and populations, along with the comparison of data across different demographics, climates, and healthcare systems. Incorporating inflammatory cytokines, viral load measurements, and markers of endothelial functions could provide deeper insights into disease severity and help in the early diagnosis of the disease before it progresses to severe illness.

## Conclusion

Our research revealed that some hematological parameters fluctuate significantly throughout the dengue disease period indicating patterns of infection and immune response. During the early days of infection, the platelet count, ESR level, hematocrit level, NLR, ALT, and AST level were greatly varied among different age groups, genders, and infection types (primary or secondary). In addition, a significant difference in the changing pattern of platelet count and NLR value and consistency of hematocrit level were observed among the groups. This result could be helpful in treating dengue patients more precisely by monitoring the biomarkers. Moreover, it will help to understand recent patterns of dengue infection and host immune responses.

## Data Availability

All data produced in the present study are available upon reasonable request to the authors

## Acknowledgement

The authors would like to thank the patients who gave consent to use their data for this study and the authority of Parkview Hospital, Chattogram, Bangladesh for their consent and support during data collection.

